# *GATA6* amplification is associated with improved survival of *TP53*-mutated pancreatic cancer

**DOI:** 10.1101/2023.03.06.23286147

**Authors:** Jung-In Yang, Amber Habowski, Astrid Deschênes, Pascal Belleau, Taehoon Ha, Edward Zhou, Chris Tzanavaris, Jeff Boyd, Christopher Hollweg, Xinhua Zhu, David Tuveson, Daniel A. King

## Abstract

**Background:** Molecular profiling of pancreatic adenocarcinoma (PDAC) demonstrates that genomic and transcriptomic features are associated with prognosis and chemosensitivity. We evaluated treatment outcomes by genetic alterations in *TP53* and *GATA6* to determine the prognostic and predictive impact of co-mutations, among patients with pancreatic cancer in New York’s largest healthcare system.

**Methods:** Retrospective analysis was performed of patients at Northwell Health diagnosed with PDAC between 2014 to 2022. Surgical status was used to segregate patients into two groups: resected and unresected. *TP53* genotype and *GATA6* amplification status were compared for overall survival (OS) as measured from time of diagnosis. Additionally, patient survival by chemotherapy regimen administered was evaluated. The Kaplan–Meier method was used to determine overall survival (OS) and the Wilcoxon test was used to compare survival curves. Previously established and published patient-derived organoids [1] were used to investigate *GATA6* expression, genetic status, and chemotherapy drug sensitivity.

**Results:** Tumor mutation status was available for 128 patients. *TP53* mutations were found in 104 patients (81.3%), *GATA6* amplifications were found in 18 patients (14.0%), and 16 (12.5%) patients had mutations in both genes. Patients with *TP53* mutations had worse OS compared to the wild-type *TP53* population (n = 22) (median OS 22.4 months, 95% CI 12.5 to 41.1, vs. 44.3 months, 95% CI 24.0 to 82.0, HR 2.03, p = 0.038). Among patients with a *TP53* mutation, a survival advantage was observed in those who had a *GATA6* amplification (n=16) compared to those who did not (n=86) (median OS 25.5 months vs. 19.4 months, HR 1.82, p = 0.027). Among patients with unresected PDAC who were *TP53*-mutant, the presence of *GATA6* amplification (n=11) was associated with a substantial survival advantage compared to *GATA6* wildtype (n=52) (median OS 25.5 months, vs. 10.1 months, HR 0.35, p = 0.004). In the *TP53* mutation group, among 33 patients who received gemcitabine and nab-paclitaxel as the first-line palliative chemotherapy, patients with *GATA6* amplification (n = 8) had significantly improved survival compared to those without *GATA6* amplification (n = 25) (mean OS 23.1 months vs 9.4 months, HR 0.52, p = 0.017). However, pancreatic cancer organoids with *TP53* mutation (n=34) did not exhibit increased drug sensitivity to GnP with *GATA6* amplification.

**Conclusions:** Genetic mutations in *TP53* were associated with shorter OS than wild-type *TP53*. We found that *GATA6* amplification appeared to attenuate poor prognosis observed in *TP53*-mutant patients regardless of type of standard chemotherapy received.

## Introduction

Pancreatic ductal adenocarcinoma (PDAC) is one of the leading causes of cancer-related death, with a five-year survival rate of 11% [2]. Only 15-20% of patients are candidates for surgical resection, despite surgical resection of pancreatic cancer being the only route toward a potential cure [3]. Furthermore, 72% of patients with complete pancreatic cancer resection have a recurrence within two years, of which 77% of cases present with distant metastases [4]. Hence, systemic therapy is essential. Two combination regimens are commonly employed in fit patients: 5-FU, oxaliplatin, and irinotecan (FOLFIRINOX) and gemcitabine with nab-paclitaxel (GnP). [5, 6]. Treatment selection between these two regimens is primarily physician choice, as randomized head-to-head comparisons of these two first-line regimens are lacking. Should biomarkers exist that predict drug responsiveness, optimal treatment could be individualized.

Broad molecular characterization of PDAC has demonstrated insight into the aggressive nature of this malignancy and suggests potential biomarkers that may guide therapy. These findings include DNA mutations in common driver genes as well as transcriptomic differences that identify pancreatic cancer subgroups, such as the classical and basal subtypes, as defined by the Moffitt criteria and widely accepted among different groups [7-10]. The classical subtype histologically resembles typical adenocarcinoma with enriched *SMAD4* loss and *GATA6* amplification, whereas the basal subtype manifests with squamous features, is enriched for *TP53* mutations, and exhibits complete loss of *CDKN2A*. In addition, transcriptomic subclasses differ by prognosis, as the basal subtype is typically more aggressive and prevalent in higher-stage disease [10].

*TP53* mutations influence pancreatic cancer progression and overall survival (OS) [11]. Moreover, *TP53* mutations can exhibit loss of normal function but can also have gain-of-function (GOF) activity depending on the type of mutation, resulting in a difference in the prognosis of colorectal cancer [12, 13]. So far, several point mutations in the DNA binding domain of p53, specifically R175H, G245S, R249S, R248Q, R248W, R273H, and R282H, are reported to have GOF, resulting in cell proliferation, metastasis, resistance, and immune evasion [12, 13]. These mutations are recently highlighted in metastatic colorectal cancer for different clinical outcomes depending on the sidedness of cancer [13]; however, whether GOF in *TP53* can affect pancreatic cancer is not well known.

In contrast to the established negative prognostic impact of *TP53* mutations on survival in pancreatic cancer, controversy exists about the prognostic impact of *GATA6*. Recent studies suggest that the *GATA6* gene expression is a marker for a relatively favorable molecular subtype of pancreatic cancer [10] however *GATA6* has been traditionally known as a proto-oncogene that is clearly associated with poor prognosis in many cancers like breast, lung, esophageal, ovarian, and other cancers, and appears causative of tumor proliferation, EMT, and metastasis [14-18]. *GATA6* copy number variation (CNV) and overexpression are frequent (∼11%) in PDAC [19], and *GATA6* expression promotes *in vitro* cell proliferation possibly through WNT/ β-catenin and other signaling pathways [20-22].

, We investigated the clinical outcomes of PDAC by comparing the genetic status of *TP53* and *GATA6* by simply comparing their genetic status without any information about their expression profile. We also compared *TP53* non-GOF and GOF to determine if survival differed by the *TP53* functional subtype. Additionally, we evaluated drug sensitivity data from human pancreatic cancer organoids to determine if *GATA6* or *TP53* mutations are predictive of response to standard-of-care chemosensitivity drugs.

## Materials and Methods

### Patients

The Northwell Health Institutional Review Board (IRB) approved this retrospective genetic study and corresponding clinical information. The patient cohort was generated by extracting patients with a diagnosis of “pancreatic cancer” or “pancreatic (ductal) adenocarcinoma” from the FoundationOne Medicine reports initiated at Northwell Health between January 2014 and March 2022. We excluded patients who did not receive any disease-modifying treatment as well as those patients who did not have at least three months of follow-up data. For survival analyses, patients were stratified in two groups: *resected* and *unresected*, as surgical status has a strong impact on prognosis. The resected pancreatic cancer group included patients who received chemotherapy in the neoadjuvant setting, adjuvant setting, or both. The unresected cancer group included patients who received palliative chemotherapy and those who received neoadjuvant therapy but did not ultimately undergo surgical removal due to progression of the disease or decline of performance status. The mean follow-up period was 15.9 months (range: 3-82 months).

### Statistical analysis

Overall survival was calculated by mutation status in *TP53* and *GATA6* and co-mutation. The Kaplan–Meier method was used to determine overall survival (OS) starting at time of diagnosis to last known follow-up or death, and the Wilcoxon test was performed using GraphPad Prism version 8.0.0 for Windows. The hazard ratio (HR) was calculated with the log-rank method. An alpha of 0.05 was considered significant for statistical analyses.

### Genetic landscape

We used FoundationOne® CDx [23] to perform genetic profiling on FFPE specimens from surgical specimens and biopsies. Genetic mutations were visualized with oncoplot using Bioconductor GenVisR package v1.20.0 in R v4.0.2. [24]. The Venn diagram was created with CRAN VennDiagram package version 1.7.1. [25]. *TP53* mutations R175H, R248W, R248Q, R249S, R273H, R273L, and R282W were grouped as a gain-of-function (GOF) [24].

### Organoids DNA Sequencing Data Processing

A subset of cancer and metastatic organoids with high cellularity was selected from Tiriac et al [1] to analyze the *GATA6* copy number and mutation landscape. As our organoid biobank contains patients who occassionally have longitudinal sampling, a random selection ensured that organoids from a single collection timepointper patient was retained. The copy numbers and the cellularity were calculated using published [1] whole-exome sequencing data using Bioconductor PureCN package version 1.18.1 [26]. After looking at the complete copy number profiles of the samples, a threshold of 1.3 and 2.7 was used to delineate the cutoff for deletions and amplifications, respectively. When more than one segment was associated with a gene, the segment with the lowest value was retained to assign a potential alteration.

### Organoids RNASequencing (RNA-seq) Data Processing and Drug Screening

The *GATA6* expression of the organoid subset was analyzed using published RNA-seq data [1]. The RNA-seq reads were aligned using STAR version 2.7.9a [27] on the GENCODE transcriptome (release 39) [28] corresponding to human genome assembly GRCh38.p13. RSEM version 1.3.3 [29] was used to extract expected counts per gene. Gene expression levels were normalized with Bioconductor DESeq2 package version 1.36.0 [30] and used for analysis.

The association between *GATA6* expression and copy number levels was tested using two-sided Pearson correlation method as implemented in R stats package version 4.2.0 [31]. The scatterplot has been generated with CRAN ggplot2 package version 3.3.6 [32].

The basal/classical subtyping of the organoids was previously defined in Tiriac et al. The density graph of the *GATA6* expression according to subtype was generated with CRAN ggplot2 package version 3.3.6 and ggridges package version 0.5.3 [33].

Drug screening data previously published in Tiriac [1] was used to compare drug sensitivity with *GATA6* genetic status and expression level. The comparison of relative AUC based on *TP53* and *GATA6* genetic status was generated by GraphPad Prism version 8.0.0 for Windows.

## Results

### Patient characteristics

There were 275 patients with pancreatic cancer assessed for eligibility, among whom 128 were included for analysis (Figure 1). The median age (in years) of this cohort was 69 years. Diverse ethnic populations were represented in this cohort with self-identified race/ethnicity as follows: 65.6% of Caucasian, 14.1% of Asian, 13.3% African American, and 5.5% of Hispanic. There were 54 (42%) patients who underwent resection and 74 (58%) patients who did not, of whom all received at least one cycle of chemotherapy.

**Figure 1.**
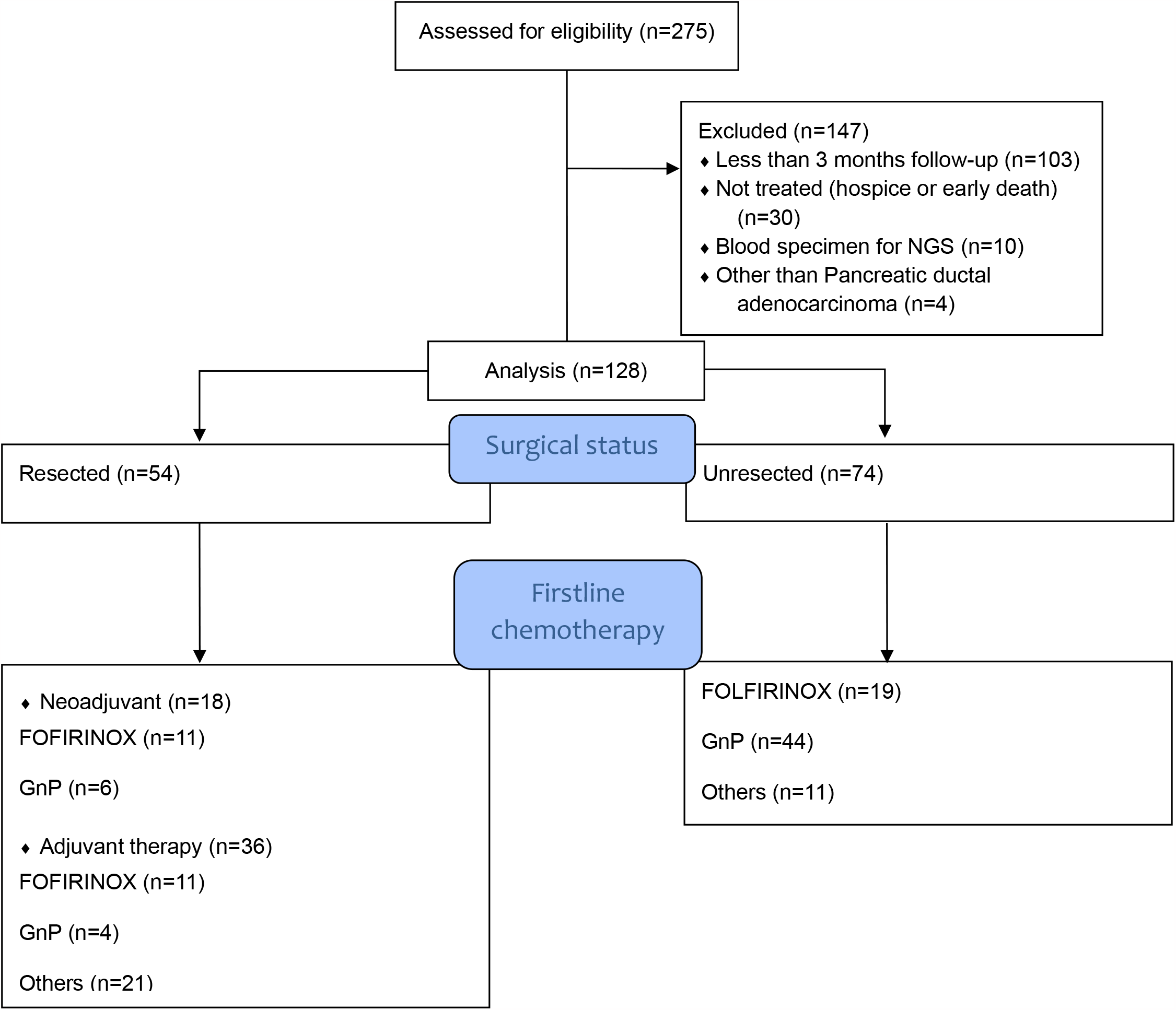
Workflow for patient selection. With exclusion criteria, 128 patients enrolled for retrospective analysis. These patients were subgrouped based on surgical status and the type of chemotherapy the patients received.

Most of the patients with palliative treatment received first-line chemotherapy (63 out of 74); 19 patients received FOLFIRINOX, and 44 patients received GnP.

### Genetic landscape

*KRAS* activation, *TP53* mutation, and *GATA6* amplifications were found in 121 (94.5%), 104 (81.3%), and 18 patients (14.0%) respectively (Figure 2a). Co-mutations in GATA6 and TP53 were found in 16 of 18 patients (Figure 2b). Regarding GOF mutations, 20 of 104 (19.2%) *TP53* mutations were GOF (Table S1).

**Figure 2.**
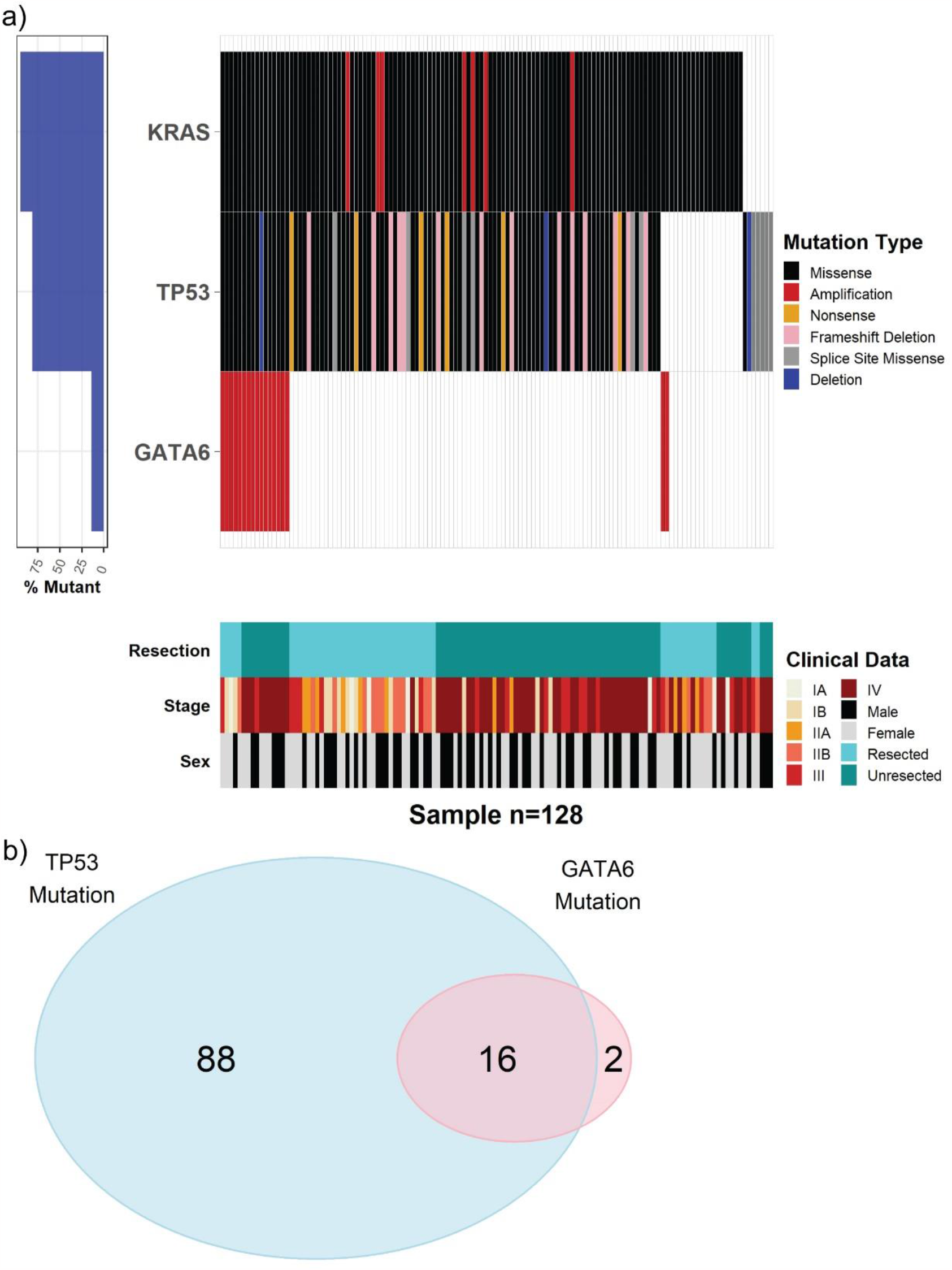
a) Mutational landscape for *KRAS, TP53*, and *GATA6* in our pancreatic cancer patients. b) Distribution of the patients for *TP53* mutation and *GATA6* amplification. The majority of our pancreatic ductal adenocarcinoma patients with *GATA6* copy number variation had a co-mutation of *TP53*.

### Clinical outcomes

#### Evaluation of TP53 subtype on survival

Patients with *TP53* mutations (n=102) had much worse OS compared to patients with wild-type *TP53* (n = 22) (median OS 22.4 months, 95% CI 12.5 to 41.1, vs. 44.3 months, 95% CI 24.0 to 82.0, HR 2.03, p = 0.038) (Figure 3a). Although the impact of *TP53* mutation in survival did not reach statistical significance when patients were divided into resected and unresected cancers, the trend of worse prognosis of TP53 persisted (Figure 3b, c.)In our cohort, GOF *TP53* mutations did not show a significant difference in OS from non-GOF *TP53*, although the trend is present in both subsets (Figure 3d).

**Figure 3.**
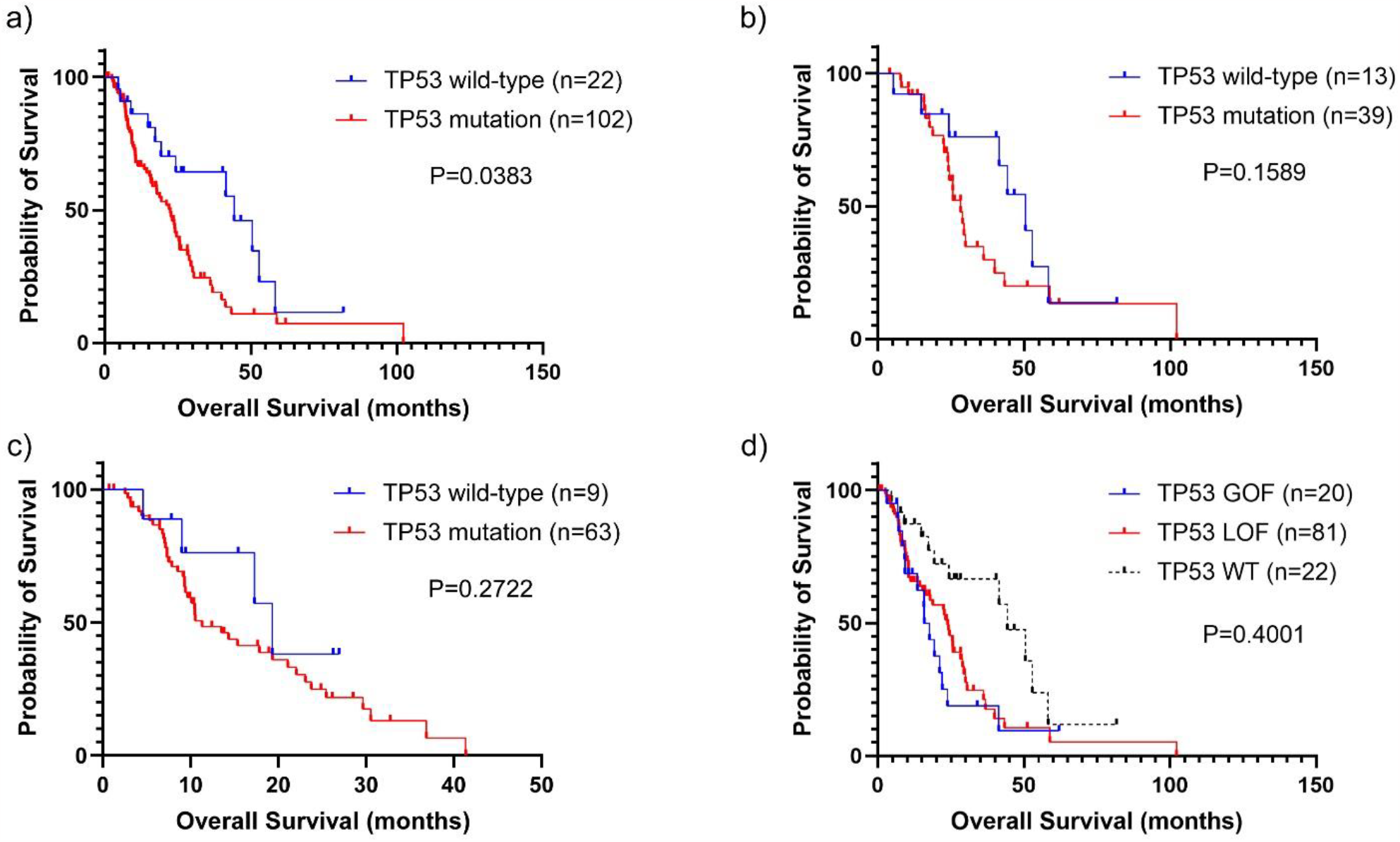
Effect of *TP53* mutation on pancreatic cancer overall survival (OS). a) Survival comparison between *TP53* mutation status only in all patients in the cohort. Patients with *TP53* mutation had a worse prognosis than the wild-type. b) Subgroup of resected pancreatic cancer was compared based on *TP53* mutation. c) Subgroup of unresected PDAC was also observed depending on *TP53* mutation. d) *TP53* gain-of-function (GOF) and loss-of-function (LOF) mutations were compared for survival analysis among pancreatic cancer patients. OS between *TP53* GOF and LOF showed no difference.

#### Evaluation of GATA6 on survival

*GATA6* amplification showed a trend toward better survival among all pancreatic cancer patients (p = 0.05) (Figure 4a). The survival difference become more apparent when focusing on the the unresected population, where the *GATA6* amplification was associated with longer survival (p=0.01) (Fig 4b). However, the majority of the *GATA6* amplified cancer patients had overlapping *TP53* mutations; therefore, we hypothesized that there might be a confounding interaction between *GATA6* and *TP53*, so we evaluated survival in the context of both genes

**Figure 4.**
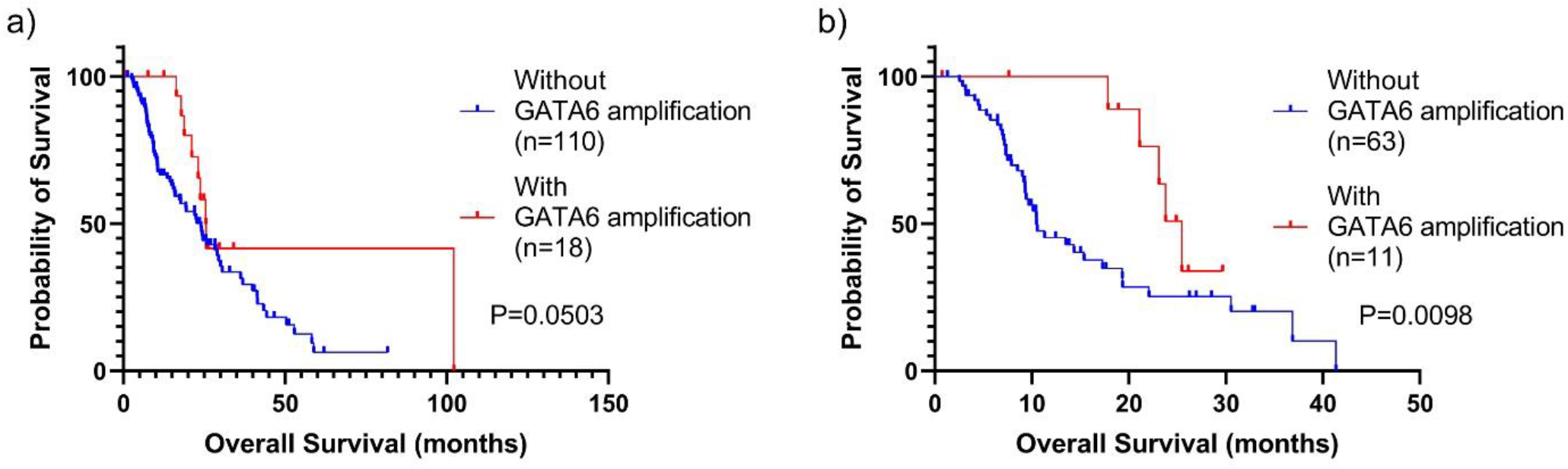
Effect of *GATA6* amplification on survival within *TP53* mutated pancreatic cancer patients. a) Overall survival (OS) was compared in all pancreatic cancer patients based on *GATA6* amplification only. Statistical significance was barely reached (p = 0.0503). b) OS in unresected subgroup based on *GATA6* status showed significant survival benefit (p = 0.0098) in pancreatic cancer with *GATA6* amplification.

First, we compared survival among *TP53*-mutant patients by *GATA6* amplification status. Among patients with a *TP53* mutation, a significant survival advantage was observed in those who had a *GATA6* amplification compared to those who did not (median OS 25.5 months, 95% CI 12.6 to 51.5, vs. 19.4 months, 95% CI 9.6 to 39.1, HR 1.82, p = 0.027) (Figure 5a), a finding that was accentuated among patients who were unresected (median OS 25.5 months, 95% CI 10.0 to 64.9 vs. 10.1 months, 95% CI 4.0 to 25.8, HR 0.35, p = 0.004) (Figure 5b). Moreover, when TP53 wild-type patients and TP53 mutation with GATA6 amplified patients were compared, their difference of survival outcomes disappeared (Figure 5c).

**Figure 5.**
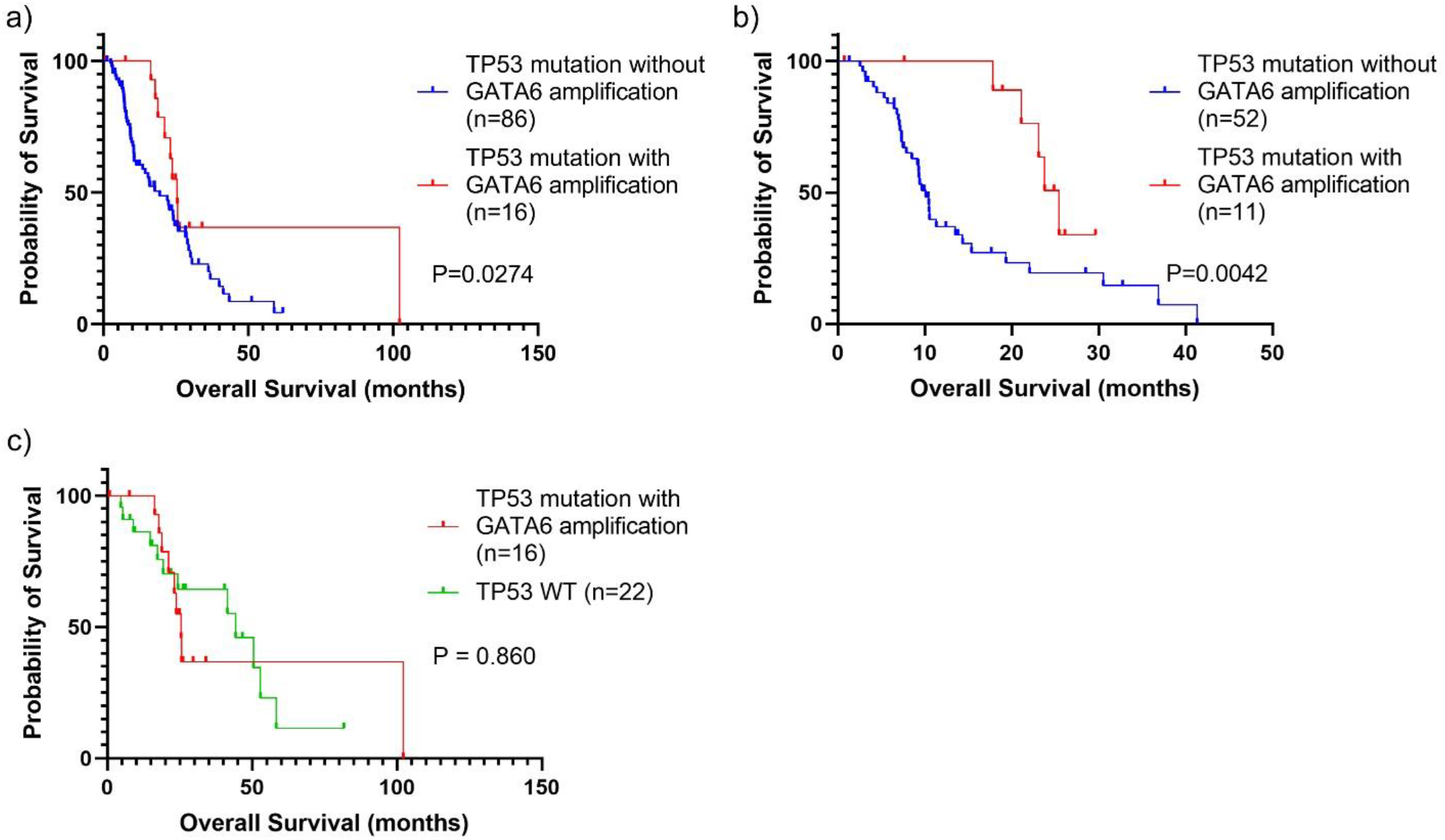
Effect of TP53 mutation and GATA6 amplification in patient survival. a) Selected patients who bear *TP53* mutation were compared based on *GATA6* amplification. Improved statistical significance was observed in *TP53* and *GATA6* subgroup analysis (p = 0.0274). b) Unresected pancreatic cancer with *TP53* mutation with *GATA6* amplification and significant better survival than *TP53* mutation without *GATA6* amplification (p = 0.0042). c) *TP53* mutants with *GATA6* amplification patients have no difference from *TP53* wild-type patients’ survival outcomes.

#### GATA6 may predict better survival in patients exposed to GnP

Lastly, we evaluated whether co-existing mutations in *TP53* and *GATA6* predicted chemotherapy response by comparing survival outcome between patients with and without *GATA6* amplifications exposed to chemotherapy within *TP53*-mutated patients. In the *TP53* mutation group, among 33 patients who received GnP as the first-line chemotherapy, patients with a *GATA6* amplification (n=8) had a significantly improved survival compared to those without *GATA6* amplification (mean OS 23.1 months, 95% CI 9.0 to 59.0, vs. 9.4 months, 95% CI 3.7 to 24.0, HR 0.52, p = 0.017) (Figure 6). The sensitivity of FOLFIRINOX could not be assessed as only one patient with *GATA6* amplification received FOLFIRINOX in our cohort.

**Figure 6.**
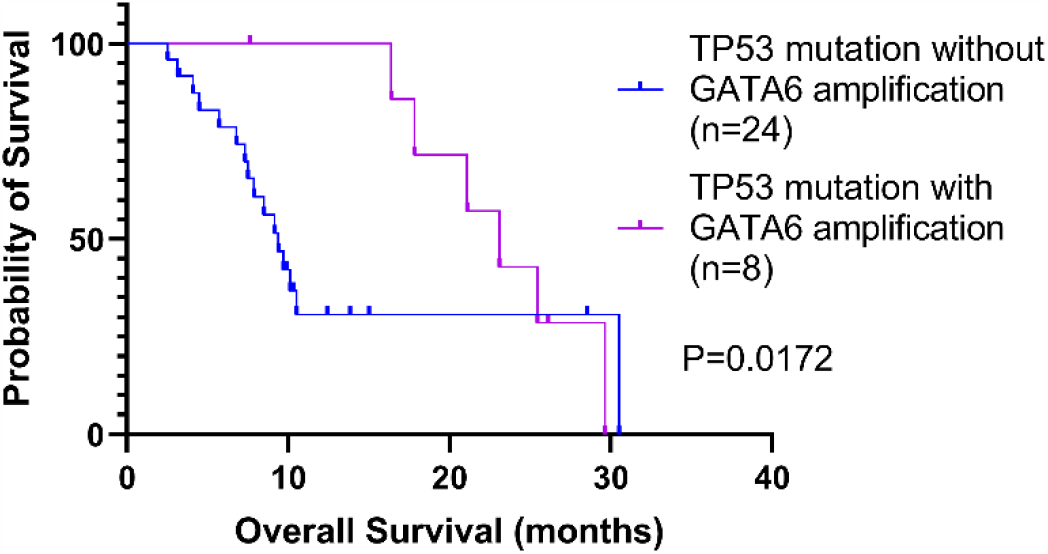
Effect of *GATA6* amplification within *TP53* mutated patients who received gemcitabine and nab-paclitaxel (GnP). Patients with pancreatic cancer-bearing TP53 mutation and GATA6 amplification at the same time had better survival benefits with GnP as first-line chemotherapy than with TP53 mutation without GATA6 amplification.

### Organoids Characteristics and drug sensitivity

Next, we investigated whether organoid data recapitulated the favorable survival signal observed in *TP53* mutant/*GATA6* mutant patients following exposure to GnP. We evaluated 37 phamacotyped organoid models that were available from Tiriac et al [1]. The *GATA6* expression levels of these organoids correlated with classical subtype, a finding previously reported by other groups [10] (Figure 7). *GATA6* amplification status had a good correlation with RNA expression level among selected organoids (Figure S2.) Among organoids, majority were *TP53* mutated and only three were*TP53* wild-type organoids. Among *TP53*-mutated organoids, 11 had *GATA6* amplification, and three of them had *GATA6* deletion (Table 1).

**Figure 7.**
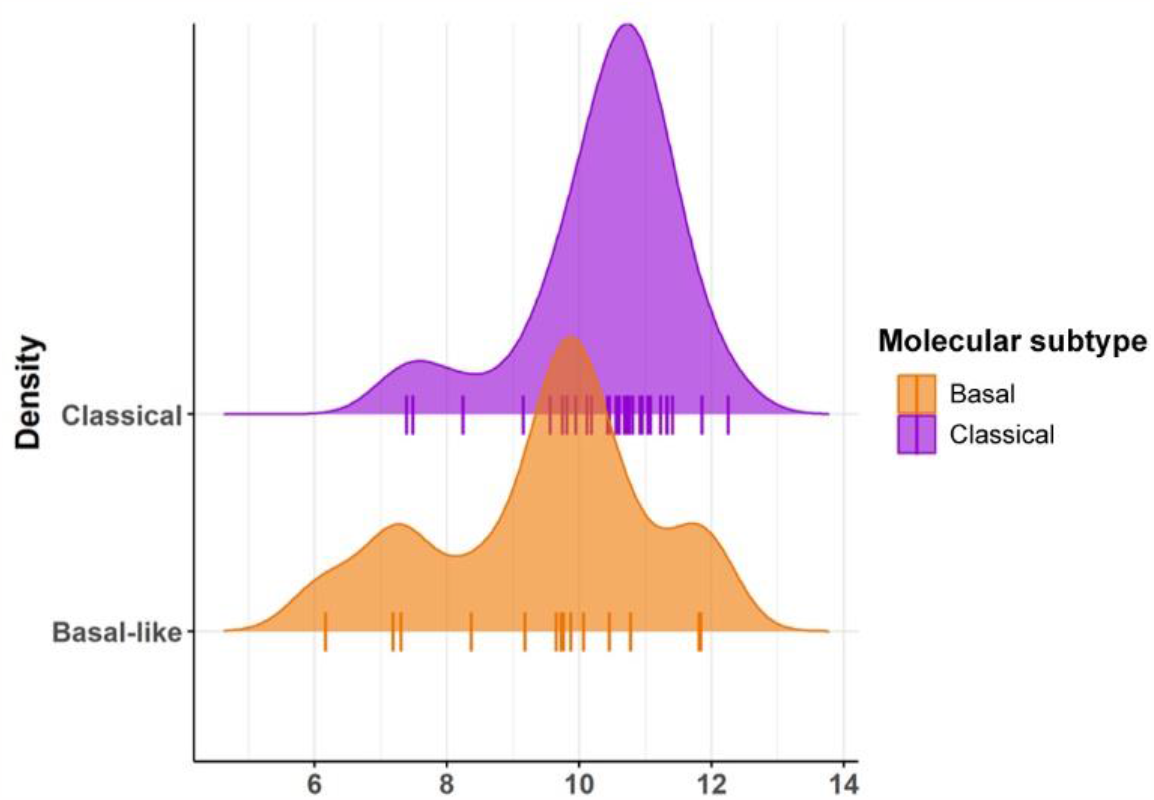
*GATA6* expression of human PDAC organoids. 37 samples of organoid have a different distribution of *GATA6* expression between classical and basal molecular subtypes.

**Table 1.** TP53, and GATA6 status in PDA organoids

| Gene | Wild-type | Mutation/Deletion | Amplification |
| --- | --- | --- | --- |
| TP53 | 3 | 34 | - |
| GATA6 | 23 | 3 (del) | 11 |

In organoids, neither *GATA6* amplification status (Figure 8a) nor its expression level (Figure 7b) had a statistically significant correlation with the drug sensitivity of five different agents, gemcitabine, paclitaxel, 5-FU, SN-38, and oxaliplatin. The relative AUC means of each drug trends towards lower AUC in *GATA6* amplification, although this is not significant. (Figure 8a), however, significant overlapping individual values are observed between GATA6 amplification and neutral groups. There was a tendency of *GATA6* over-expressed organoids to be more susceptible to oxaliplatin and 5-FU than *GATA6* low organoids (Figure 8b), however, the statistical difference was not significant. In contrast to the clinical observation, neither gemcitabine nor paclitaxel predicted chemo-sensitivity to *GATA6* amplified and *TP53* mutated organoids.

**Figure 8.**
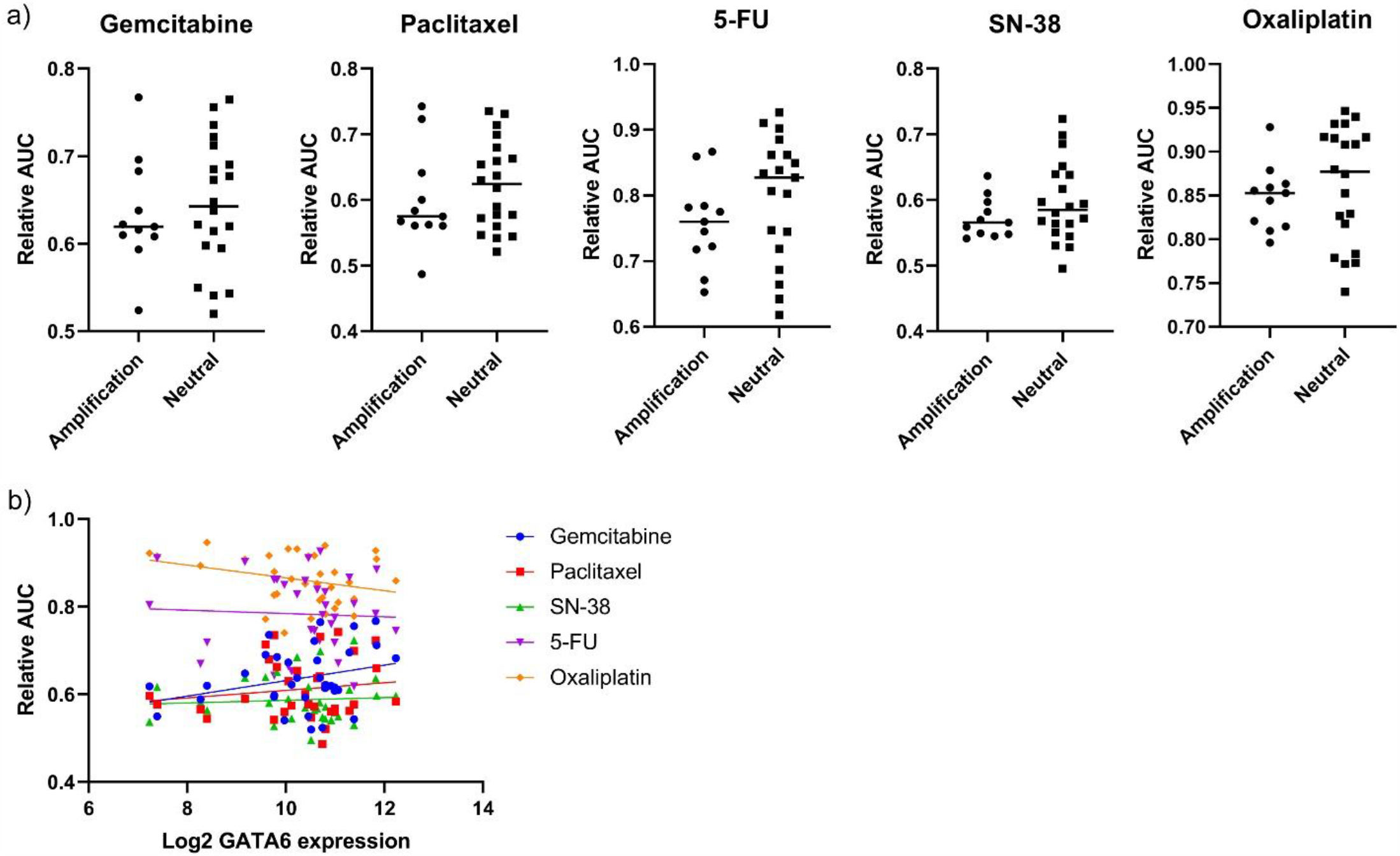
Drug sensitivity of human PDAC organoids with *TP53* mutations based on *GATA6* status. a) *GATA6* CNV has no statistical power to show chemosensitivity to conventional single chemo-reagent, such as gemcitabine, paclitaxel, 5-FU, SN38, and oxaliplatin. Bar showing mean of relative AUC. b) Higher *GATA6* expression level also failed to show a better response to reagents. Furthermore, each linear graph displays a flattened slope representing no correlation between the GATA6 expression level and relative AUC.

## Discussion

We have observed that previously reported poor prognosis associated with *TP53* mutation [11] and favorable prognosis of *GATA6* amplification [10] were reproduced in our retrospective cohort. Our analysis showed *GATA6* amplification had clear benefit of survival benefit in PDAC contrary to negative expectation in different solid cancers.

Most of our patients’ PDAC with *GATA6* amplification had co-mutation of *TP53*, hence we looked at subgroup of patients within *TP53* mutated PDAC. Our study suggested that *GATA6* amplification may partially compensate for an unfavorable consequence of *TP53* mutation in PDAC especially in a palliative setting. Response to systemic chemotherapy in the co-existence of *GATA6* amplification and *TP53* mutation was more dramatic than *TP53* mutation alone in our patient data but was not observed in our organoid model. We did not observe statistically significant increased sensitivity to a specific chemotherapy associated with *GATA6* amplification among *TP53* mutant human pancreatic cancer organoids from Tiriac et al.

While PDAC organoids better recapitulate cancer cell biology compared to 2D cell lines, they do not encompass all the cell types in the tumor. As we did not see increased sensitivity of *GATA6* overexpressing organoids towards chemotherapy, while we did see *GATA6* amplified tumors were more sensitive to GnP, this may suggest the increased sensitivity *in vivo* may be due to how the chemotherapy influences host-tumor interaction, and not cancer cell viability alone. Additionally, organoid drug screening was performed using monotherapy, not combination cocktails as given to patients. The impact of combination therapy on organoid response should be explored in future work However, the data from human pancreatic cancer organoids could be misleading due to their simplified nature that is imperfectly capitulating in vivo physiology. Human organoids were subjected to only a single drug, not a combination of chemotherapy, such as gemcitabine plus nab-paclitaxel. There is uncertainty if two drugs can interact and change the trend of drug screening. Moreover, the organoid model does not account for TME which could largely influence the drug’s metabolism that may change the relative AUC.

Nevertheless, experimental evidence did not support a predictive relationship between chemotherapy and GATA6. This suggests that *GATA6* amplification could be a marker of prognosis from the nature of cancer itself rather than a predictive marker of specific chemotherapy.

A possible mechanism of *GATA6* in pancreatic cancer could be its ability to differentiate the cancer cells from the dedifferentiated state and EMT effect caused by *KRAS* and *TP53* mutations. One study showed that *GATA6* ablation resulted in the upregulation of the EGFR pathway mediated by constitutively active *KRAS* (G12V) [34]. *GATA6* also inhibits EMT in vitro and cell dissemination in vivo, possibly through *ΔNp63* expression or *TP53* mutation [35, 36].

*GATA6* gene expression was an essential gene determining classical subtype or basal subtype, and *GATA6* was a potential predictor of chemosensitivity in the COMPASS trial [10, 37]. *GATA6* may partly play a role in turning on and off the switch for classical subtype and basal subtype involving epigenetic mechanism. For example, one study demonstrated inhibiting a suppressive epigenetic modulating enzyme, EZH2, could induce *GATA6* expression in basal subtypes and trigger conversion to a classical subtype [38]. In terms of evaluating chemosensitivity associated with *GATA6*, our organoid data suggested that the gene had some increased sensitivity to oxaliplatin, although was not statistically significant.

In this cohort, we did not have an adequate sample size to analyze the clinical outcome from wild-type *KRAS* with *GATA6* amplification. It was previously shown that both *KRAS* and *TP53* mutations are independently associated with poor prognosis [39], and our cohort with *TP53* mutation co-harbored *KRAS* mutation at the same time. Therefore, it will be interesting to see the interaction between *GATA6* amplification with *KRAS* activation.

Further investigation of the underlying mechanism involving *GATA6* in PDAC will broaden our understanding of the disease and hopefully improve survival rates of pancreatic cancer patients by identifying genetic markers of outcomes and ideal therapies. Additionally, combining drugs targeting epigenetic modulators for pancreatic cancer systemic therapy could be an emerging therapeutic option for pancreatic cancer.

## Conclusion

Our single institution exploratory study using NGS showed that *TP53* status and *GATA6* amplification are significantly associated with clinical outcomes of pancreatic cancer. More detailed clinical implications of *GATA6* amplification should be assessed in future studies. Also, further investigation of the underlying mechanism with *GATA6* in PDAC will broaden our understanding of the disease and find a way to improve sensitivity to systemic therapy.

## Supporting information

Supplement_patients

## Data Availability

All data produced in the present study are available upon reasonable request to the authors
All data produced in the present work are contained in the manuscript

## An Author Contribution statement

J.Y. prepared manuscript and collected data; T.H. and C.H. provided statistical analyses; C.H., E.Z., and C.T. collected patient clinical data; A.H., A.D., and P.B conducted genetic analyses and organoid data processing; J.B. provided statistical analyses strategy; X.Z. provided patient dataset with NGS; D.T revised manuscript; D.K. oversaw the project and is the corresponding author.

---

**Figure S 1.**
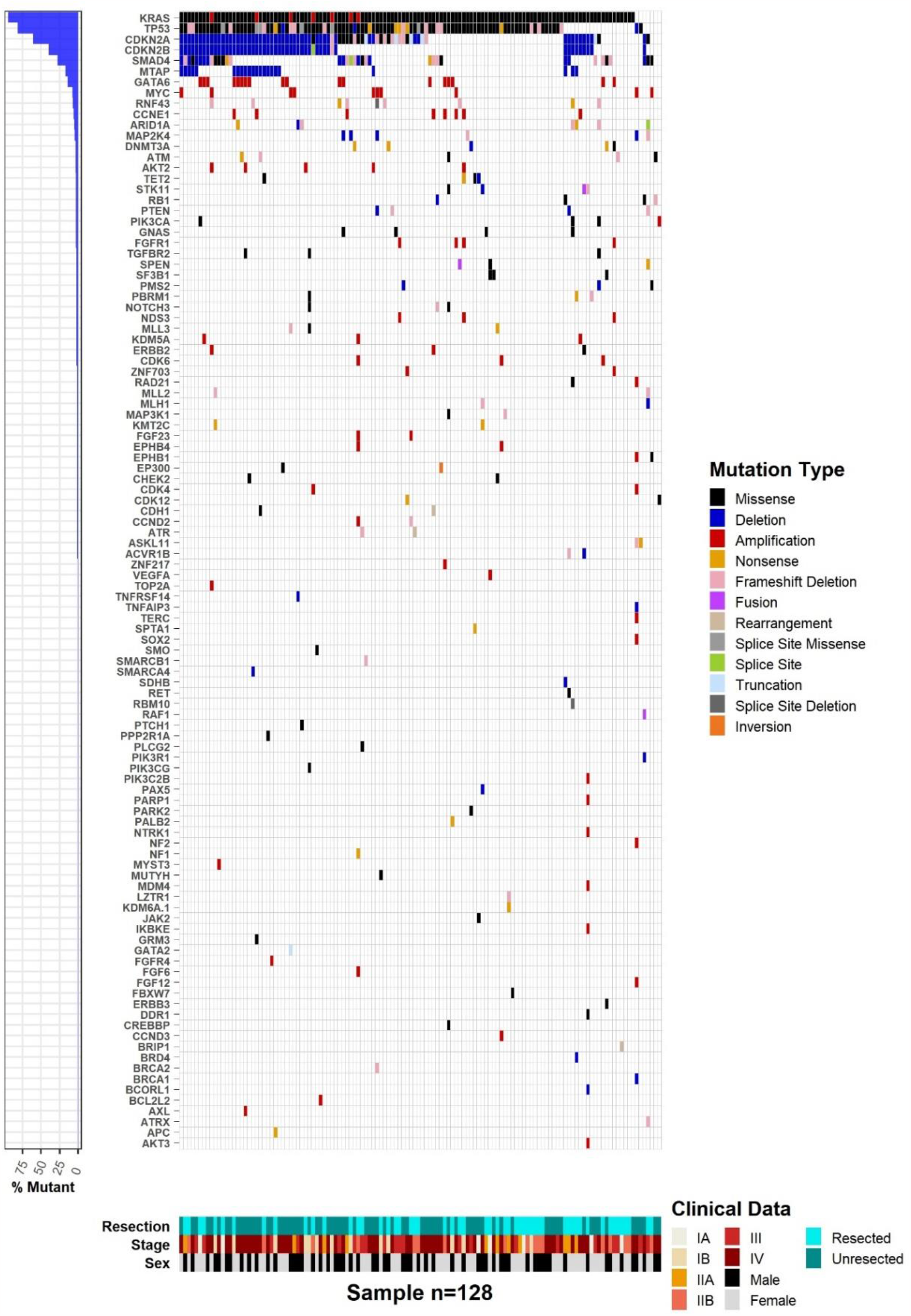
Mutational landscape for all genetic changes in genes tested by FoundationOne

**Figure S 2.**
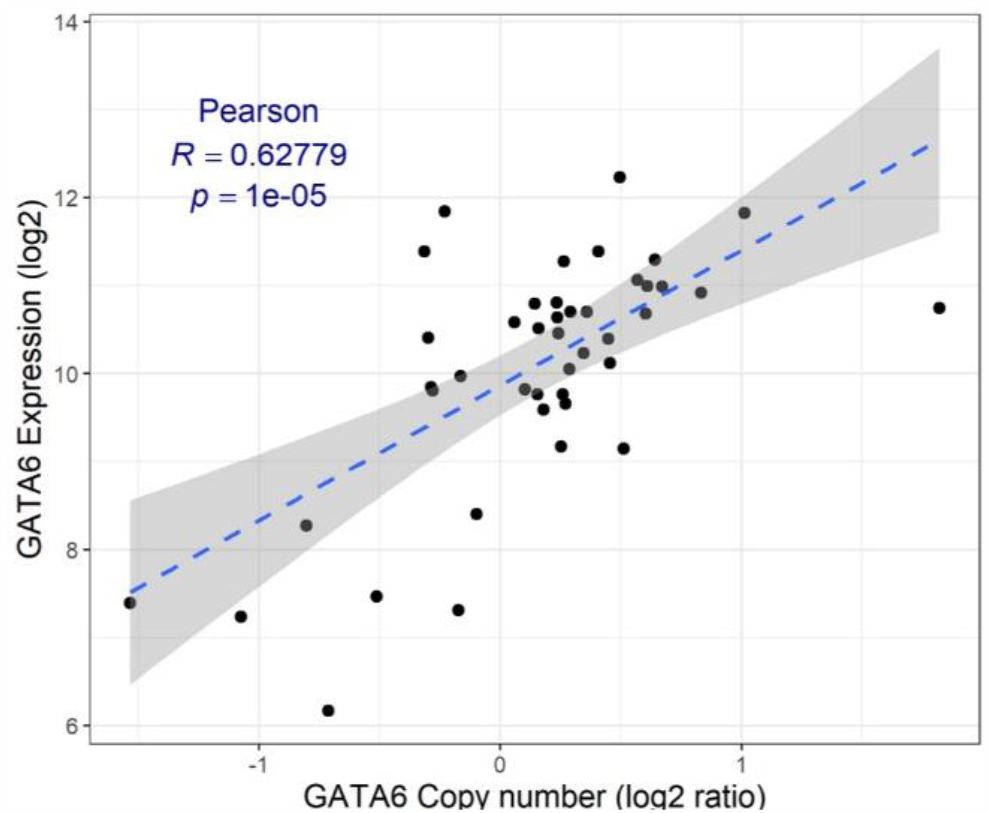
The correlation between *GATA6* CNV and *GATA6* mRNA expression. The amount of *GATA6* CNV is proportional to the *GATA6* expression level

**Table S1.** Gain-of-function *TP53* distribution (n = 20)

| Point mutation | Number of patients |
| --- | --- |
| R175H | 8 |
| R248Q | 5 |
| R273H | 4 |
| R282H | 2 |
| R248W | 1 |
| R249S | 0 |
| G245S | 0 |

